# Community engagement and involvement in national perioperative quality improvement: a mixed-methods assessment of knowledge, attitudes, and practices among stakeholders in Ethiopia

**DOI:** 10.64898/2026.11.24.26368817

**Authors:** Kokeb Desta, Iverson Katherine, Tesfay Yohannes Ambese, Kelem Desta, Bezaye Zemedkun, Mohammed Aman, Nuroadis Guchima, Bethelhem Mulye, Tigist Fantahun, Bruce Biccard, Fitsum K Belachew

**Affiliations:** Department of Anesthesia, School of Medicine, Debre Berhan University; N4PCc CEI Lead, Ethiopia; Network for Perioperative and Critical Care (N4PCc), Debre Berhan University; Asrat Woldeyes Health Sciences Campus, Ethiopia; Division of Trauma, Surgical Critical Care, Burns and Acute Care Surgery, Department of Surgery, University of California San Diego, San Diego, CA, USA; Department of Statistical and Data Governance, Network for Perioperative and Critical Care (N4PCc), Addis Ababa, Ethiopia; Department of Midwifery, School of Nursing and Midwifery, Debre Berhan University, Ethiopia; Department of Anesthesia, School of Medicine, Menelik Second Medical College, Addis Ababa, Ethiopia; Department of Public Health, Debre Birhan Health Centre, Tena Extension Worker Coordinator, N4PCc CEI patient researcher; School of Medicine, SPMMC; National Perioperative Quality Improvement Network (NaPQIN) Quality Officer, N4PCc, Ethiopia; Department of Nursing, Hakim Gizaw Hospital; N4PCc Patient Researcher, Ethiopia; Department of Clinical Neurosciences, University of Oxford, Oxford, UK

## Abstract

**Background:** Sustainable implementation of perioperative quality registries in low- and middle-income countries requires more than technical infrastructure; it also requires informed stakeholders, supportive attitudes, and embedded practice. Ethiopia’s National Perioperative Quality Improvement Network (NaPQIN), established across 10 hospitals, provided an opportunity to examine whether structured community engagement and involvement (CEI) could improve knowledge, attitudes, and practices among healthcare stakeholders.

**Methods and findings:** We conducted a convergent parallel mixed-methods study from July 2024 to December 2025 across 10 NaPQIN hospitals in Ethiopia. Pre-intervention (n=25) and post-intervention (n=18) KAP surveys were administered around a three-month CEI program that included training, information materials, workshops, supervision, and stakeholder engagement. Semi-structured interviews (n=14) provided qualitative context. Quantitative data were analyzed descriptively, qualitative data were analyzed thematically, and findings were integrated through triangulation. Knowledge improved across most domains, with the largest gains in identifying the primary purposes of a surgical registry (+12.0 percentage points) and non-characteristics of a registry (+10.4 percentage points). Attitudes improved across all measured domains, with the largest gains in overall satisfaction with NaPQIN (+0.60 on a 5-point scale), perceived CEI contribution to NaPQIN sustainability (+0.59), and the importance of community engagement (+0.44). Practices improved more modestly: clinician daily registry use increased by 21.3 percentage points and institutional CEI formalization by 20.4 percentage points, whereas benchmarking use declined by 8.9 percentage points. Qualitative findings highlighted the value of perioperative data, persistent implementation barriers, and practical strategies for strengthening registry use in low-resource settings.

**Conclusions:** CEI was associated with improved stakeholder knowledge and attitudes toward NaPQIN, with early signs of practice integration. These findings suggest that CEI may be an important component of perioperative quality improvement networks in low- and middle-income country settings, while also highlighting ongoing needs in workflow support, benchmarking capacity, and digital infrastructure.

## Introduction

Surgical conditions account for an estimated 30% of the global burden of disease, yet low,and middle-income countries (LMICs), where nine out of ten people cannot access safe, timely surgery, contribute only 3.5% of global surgical volume while bearing disproportionately high morbidity and mortality (1,2). A foundational barrier to improving this situation is the absence of reliable, locally owned perioperative data (3). Without systematic measurement, identifying system bottlenecks, benchmarking care quality, and designing evidence-based interventions remain aspirational rather than achievable (3).

Perioperative registries address this data deficit directly. By capturing patient-level clinical data across the perioperative pathway, they enable quality improvement (QI) auditing, complication monitoring, resource allocation, and national policy development (3). Despite their demonstrated value in high-income countries, perioperative registries remain rare across sub-Saharan Africa, where paper-based systems, fragmented data flows, workforce constraints, and limited digital infrastructure conspire against implementation (3–5). A systematic review by Kifle et al. (2023) identified that while registry implementation in LMICs is feasible, local stakeholder engagement and sustained monitoring remain the most critical determinants of success (3).

Recognizing these realities, African Perioperative Research Group (APORG) conducted a Delphi consensus study to establish a minimum standardized perioperative dataset for African clinical settings, emphasizing that electronic data capture systems must be designed with and for local clinicians to realize their potential (6). Building on this work, Ethiopia, through a collaboration between the Network for Perioperative and Critical Care (N4PCc), the NIHR Global Health Research Unit - African Partnership for Perioperative and Critical Care Research (APPRISE), established the National Perioperative Quality Improvement Network (NaPQIN) in June 2023. NaPQIN now encompasses more than 10 hospitals across referral, general, and primary care facility levels in multiple Ethiopian regions and constitutes a landmark LMIC initiative in real-time perioperative data capture (5).

However, establishing a registry is not the same as embedding it sustainably into clinical culture. The World Health Organization (WHO) defines community engagement as a process of developing relationships that enable stakeholders to work together to address health-related issues and promote well-being (7). Evidence consistently shows that community engagement and involvement (CEI), encompassing stakeholder orientation, patient-researcher participation, training, and iterative feedback, are essential facilitators of health system program sustainability in African contexts (8,9). Without deliberate CEI strategies, even technically sophisticated registries risk poor uptake, incomplete data, and eventual abandonment (9).

Despite this recognition, empirically evaluated CEI interventions specifically targeting perioperative registry stakeholders in LMIC settings are absent from the published literature. This paper addresses that gap. We present the design, implementation, and evaluation of a structured CEI program across NaPQIN’s ten member hospitals, assessing its effects on stakeholders’ Knowledge, Attitude and Practice (KAP) through a convergent parallel mixed-methods study. Our findings offer practical, transferable lessons for LMIC perioperative quality networks seeking to build the stakeholder engagement infrastructure on which lasting improvement depends.

## Materials and methods

### Ethics statement

The study was approved by the Debre Berhan University Asrat Woldeyes Health Science Campus Institutional Review Board (Protocol No. IRB-095) before study commencement. All participants gave written and verbal informed consent before taking part. For the Google Forms surveys, we obtained consent on the first page of the form; for the interviews, we obtained consent before the interview began, including consent to audio recording. Participation was voluntary, and participants could withdraw at any time.

#### Study design

We used a convergent parallel mixed-methods design, combining a pre-post quantitative KAP survey with qualitative semi-structured stakeholder interviews. Quantitative and qualitative data were collected concurrently, analyzed independently, and integrated during interpretation to provide comprehensive explanatory insight into NaPQIN implementation. The study was conducted from July 2024 to December 2025 in collaboration with N4PCc, patient researchers, and NaPQIN member hospitals across Ethiopia. The quantitative component is reported in accordance with the Strengthening the Reporting of Observational Studies (STROBE) guidelines for observational studies, and the qualitative interview component is reported in accordance with the Consolidated Criteria for Reporting Qualitative Research (COREQ) checklist. Completed reporting checklists will be provided as supporting information at submission.

The study was conducted in ten NaPQIN member hospitals, spanning referral, general, and primary facilities across the Amhara and Addis Ababa regions and beyond. Purposive and convenience sampling were used to recruit key stakeholders who were actively involved in NaPQIN-related activities during the study period.

The quantitative component enrolled clinicians (surgeons, anesthesiologists, general practitioners, nurses, gynecologists), hospital leaders (chief clinical directors, OR directors, surgical department heads), site supervisors, data collectors, and QI coordinators. The qualitative component engaged 14 participants, including data collectors (n=3), surgeons (n=3), administrators and QI officers (n=3), hospital supervisors (n=2), and Ministry of Health officials (n=3), with experience ranging from several months to more than 8 years.

#### Pre- and post-intervention KAP assessment

A structured questionnaire covering knowledge, attitudes, and practices regarding perioperative registries and NaPQIN was developed by the CEI and patient researcher team, validated by registry experts, and administered electronically via Google Forms. Pre-assessment was conducted at baseline (July-August 2024), followed by three months of CEI activities, and then post-assessment approximately three months after the intervention concluded.

#### CEI intervention and capacity building

Interventions were co-designed with CEI principles and input from patient researchers to foster stakeholder ownership and sustainability. Activities included:

- Orientation and training sessions (in-person and virtual via Zoom) for hospital leaders, supervisors, and data collectors, covering CEI roles, perioperative registry importance, NaPQIN concepts, QI strategies, and data management principles
- Distribution of Information, Education, and Communication (IEC) materials, including brochures summarizing training content
- Face-to-face small-group discussions and CEI-led stakeholder engagement workshops at selected hospitals
- Supervision, site visits, and structured interviews by the CEI team (including patient researchers) to explore challenges, needs, and solutions
- Targeted reinforcement of registry use, benchmarking practices, and integration of data capture into routine clinical workflows

#### Qualitative stakeholder interviews

Semi-structured interviews (n=14) explored contextual factors influencing NaPQIN adoption. An interview guide addressed the perceived value of perioperative data, barriers and facilitators to registry implementation, strategies for success, and recommendations for low-resource settings. Interviews were conducted by trained members of the CEI team, including patient researchers, who were involved in program implementation and had prior familiarity with some participants through NaPQIN activities. Interviews were audio-recorded with consent, conducted in English or Amharic as appropriate, and held in private locations at participating hospitals or by telephone/virtual call when in-person interviews were not feasible. Brief field notes were recorded during and after interviews to capture contextual observations. Audio recordings were transcribed verbatim, and Amharic interviews were translated into English for analysis.

#### Sample size and sampling

For the quantitative component, convenience sampling enrolled all available NaPQIN stakeholders during the implementation period (n=25 pre-intervention; n=18 post-intervention). For the qualitative component, purposive sampling was used to ensure diversity of roles and experiences (n=14).

For the quantitative component, all available responses were included in the descriptive analysis for each item. No a priori sample size calculation was performed because the study was designed as an early implementation evaluation within an existing program. For the qualitative component, sampling continued until a diverse range of stakeholder roles and implementation experiences had been captured and no major new issues were arising in subsequent interviews.

#### Data analysis

Quantitative data were cleaned and analyzed descriptively using Stata v17, reporting frequencies, percentages, means, and pre-post changes. Because the pre- and post-intervention samples were small, unmatched, and intended to provide an early implementation assessment, no formal hypothesis testing was undertaken. Missing quantitative data were handled by complete-case reporting for each item. Qualitative data underwent inductive thematic analysis of interview transcripts. Two members of the research team reviewed transcripts, iteratively developed initial codes, discussed and refined the coding framework, and grouped codes into themes through team discussion. Field notes were used to support contextual interpretation, and a brief summary of emerging findings was shared with a subset of participants to support credibility. Quantitative and qualitative findings were integrated through triangulation, with quantitative KAP results contextualized by qualitative themes.

## Results

### Participant characteristics

A total of 25 participants completed the pre-assessment, and 18 completed the post-assessment. The pre- and post-intervention samples were not matched at the individual level. The overall demographic profile was broadly similar across time points, which supports interpretation of the observed KAP changes while not eliminating the possibility of sample-related differences. The majority of participants were male at both time points (84.0% pre; 83.3% post).

Hospital chief clinical directors comprised the largest single position category (28.0% pre; 27.8% post), alongside surgical department heads and other administrative-clinical roles. Surgeons represented the largest professional group (28.0% pre; 33.3% post), followed by general practitioners, nurses, public health professionals, gynecologists, and anesthesiologists. Most participants had over five years of professional experience (76.0% pre; 72.2% post). Full sociodemographic data are presented in **Table 1**.

**Table 1.**
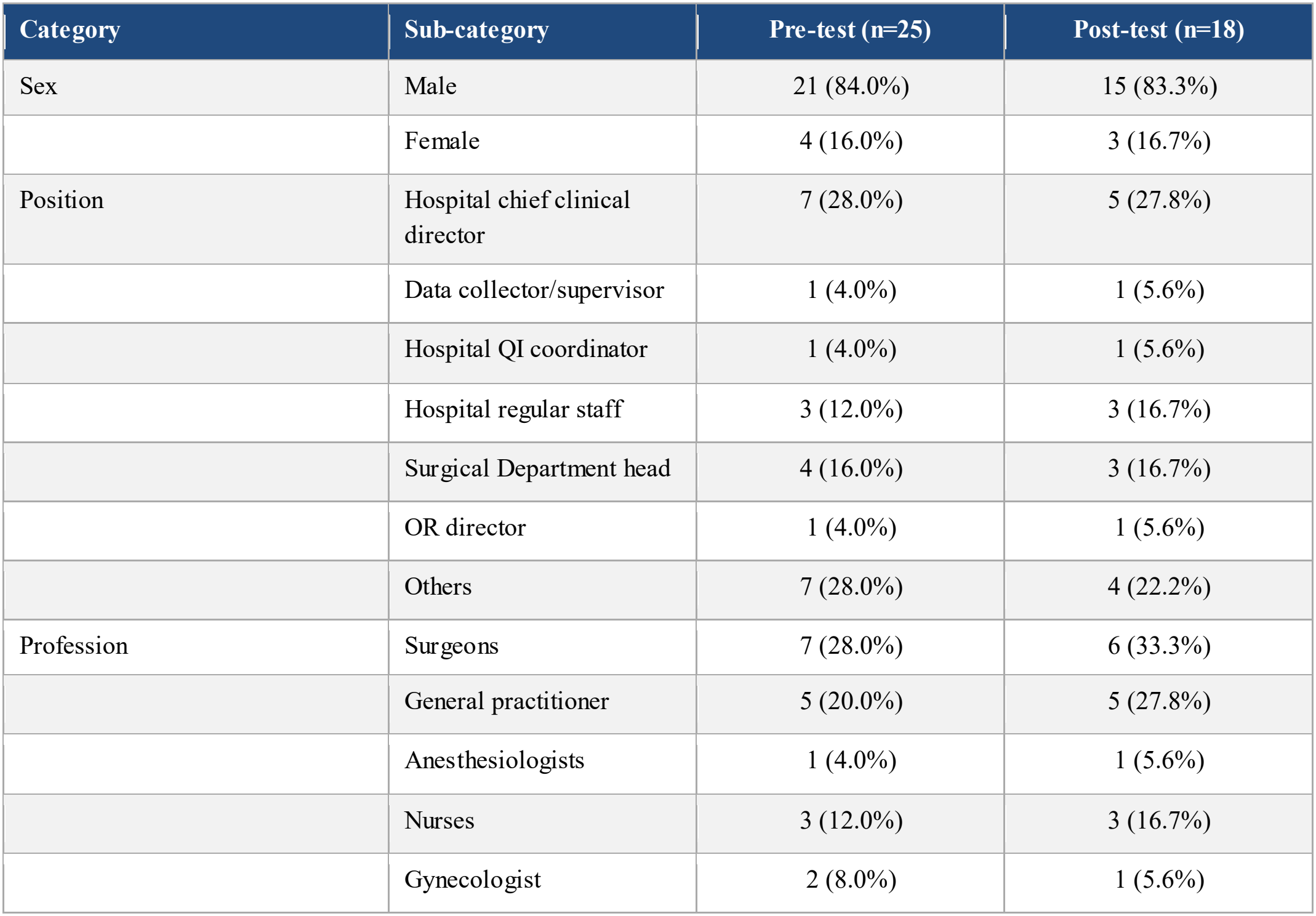
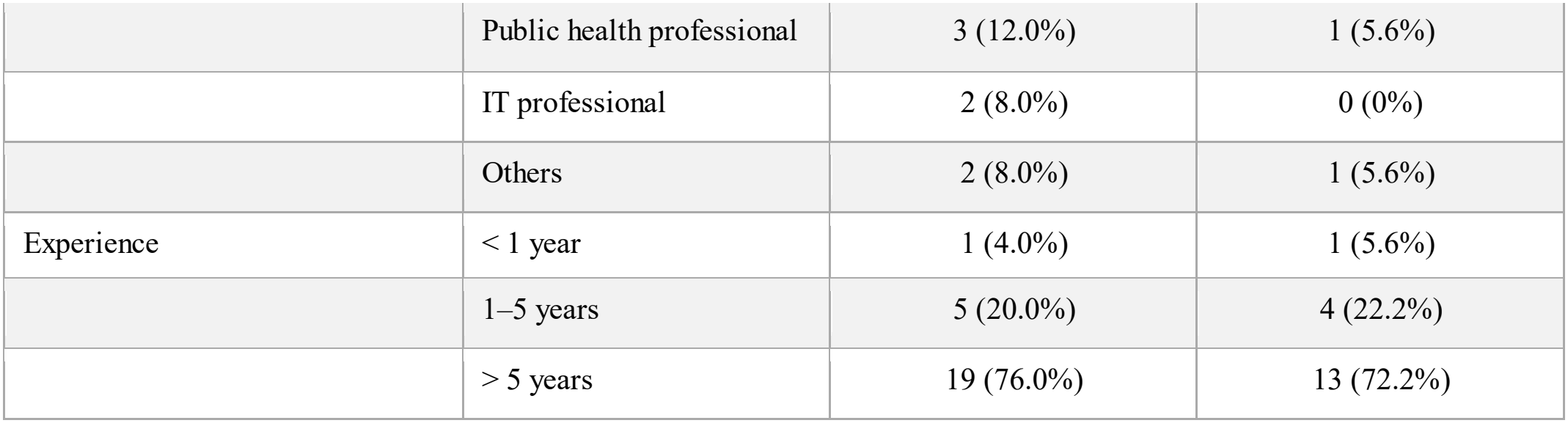
Sociodemographic characteristics of study participants.

### Knowledge assessment

Post-intervention knowledge improved in six of eight assessed domains (**Table 2**). The most substantial gain was in identifying the primary purposes of a surgical registry (+12.0 percentage points(pp)), rising from 88% to 100% correct. Correct identification of non-characteristics of a perioperative registry also increased markedly (+10.4 pp). Awareness of eligible NaPQIN member institutions and the non-objective of NaPQIN each increased by 6.4 pp. Knowledge of NaPQIN membership benefits remained at 100% at both timepoints.

**Table 2.** Pre- and post-intervention knowledge assessment results.

| Knowledge Domain | Pre-test (% Correct) | Post-test (% Correct) | Δ Change |
|---|---|---|---|
| Identifying the non-controlling purpose of NaPQIN | 80% (20/25) | 88.9% (16/18) | **+8.9%** |
| Identifying non-methods of NaPQIN advocacy | 92% (23/25) | 88.9% (16/18) | −3.1% |
| Benefits of NaPQIN membership | 100% (25/25) | 100% (18/18) | 0% |
| Eligible NaPQIN member institutions | 88% (22/25) | 94.4% (17/18) | **+6.4%** |
| Comprehensive NaPQIN goals (incl. SaLTS) | 100% (25/25) | 94.4% (17/18) | −5.6% |
| Non-objective of NaPQIN | 88% (22/25) | 94.4% (17/18) | **+6.4%** |
| Primary purposes of a surgical registry | 88% (22/25) | 100% (18/18) | **+12.0%** |
| Non-characteristics of a perioperative registry | 84% (21/25) | 94.4% (17/18) | **+10.4%** |

Two domains declined slightly: recognition of non-methods of NaPQIN advocacy (−3.1 pp) and comprehensive NaPQIN goals, including SaLTS implementation (−5.6 pp).

### Attitude assessment

Post-intervention attitudes shifted positively across all 13 assessed dimensions (**Table 3**). The largest increases were observed in overall satisfaction with NaPQIN’s registry efforts (+0.60), perceived CEI contribution to NaPQIN-specific sustainability (+0.59), and perceived CEI contribution to quality improvement projects in general (+0.58).

**Table 3.** Pre- and post-intervention attitude assessment results (5-point Likert scale)

| **Attitude Statement (5-point Likert scale)** | **Pre-test Mean** | **Post-test Mean** | **Δ Change** |
|---|---|---|---|
| Sustainability of NaPQIN hospital participation | 4.52 | 4.72 | +0.20 |
| NaPQIN enhances inter-hospital surgical collaboration | 4.56 | 4.89 | +0.33 |
| Registry cannot be integrated into workflow (reverse-scored) | 2.00 | 2.39 | +0.39 |
| Registries improve leadership decision-making | 4.68 | 4.83 | +0.17 |
| Registries support reporting to health authorities | 4.56 | 4.72 | +0.16 |
| Registries enable evidence-based intervention planning | 4.52 | 4.78 | +0.36 |
| Registries allow effortless quality tracking | 4.36 | 4.61 | +0.25 |
| Registries upgrade skills through evidence-based learning | 4.44 | 4.78 | +0.34 |
| Community engagement is essential for QI | 4.28 | 4.72 | +0.44 |
| Overall satisfaction with NaPQIN registry efforts | 3.84 | 4.44 | +0.60 |
| Patient feedback improves the quality of care | 4.44 | 4.83 | +0.39 |
| CEI increases ownership & sustainability in general QI | 4.20 | 4.78 | +0.58 |
| CEI increases ownership & sustainability in NaPQIN specifically | 4.24 | 4.83 | +0.59 |

Other increases included the perceived importance of community engagement for quality improvement (+0.44), the belief that patient feedback improves care quality (+0.39), the perceived value of registries for evidence-based intervention planning (+0.36), perceptions of NaPQIN’s role in enhancing surgical collaboration (+0.33), and its educational value for skills upgrading (+0.34).

The reverse-scored item, “Registry cannot be integrated into clinical workflow,” increased from 2.00 to 2.39 (+0.39).

### Practice assessment

Practice-level changes were more modest than changes in knowledge and attitudes (**Table 4**).

**Table 4.** Pre- and post-intervention practice assessment results.

| **Practice Indicator** | **Pre-test (% Yes)** | **Post-test (% Yes)** | **Δ Change** |
|---|---|---|---|
| Hospital is a NaPQIN member | 40% (10/25) | 50.0% (9/18) | +10.0% |
| Hospital implements a surgical perioperative registry | 28% (7/25) | 38.9% (7/18) | +10.9% |
| Registry used for institutional reporting | 32% (8/25) | 39.0% (7/18) | +7.0% |
| Clinicians use registry for daily clinical activities | 12% (3/25) | 33.3% (6/18) | +21.3% |
| Leadership uses registry data for decision-making | 20% (5/25) | 33.3% (6/18) | +13.3% |
| NaPQIN used for inter-institutional benchmarking | 20% (5/25) | 11.1% (2/18) | −8.9% |
| Registry data used for quality improvement projects | 20% (5/25) | 22.2% (4/18) | +2.2% |
| Institution has formalized CEI practice in QI projects | 24% (6/25) | 44.4% (8/18) | +20.4% |

Clinicians’ daily registry use increased from 12.0% to 33.3% (+21.3 pp). Institutional formalization of CEI practices in quality improvement projects increased from 24.0% to 44.4% (+20.4 pp). Leadership use of registry data for decision-making increased from 20.0% to 33.3% (+13.3 pp), and hospital-level surgical registry implementation increased from 28.0% to 38.9% (+10.9 pp).

NaPQIN benchmarking use declined from 20.0% to 11.1% (−8.9 pp). Use of registry data for formal quality improvement projects increased from 20.0% to 22.2% (+2.2 pp). Only 40% (10/25) of participants before the intervention and 50.0% (9/18) after it reported that their hospital is a NaPQIN member, although all were recruited from NaPQIN hospitals (Table 4).

### Qualitative findings

Fourteen semi-structured interviews yielded four major themes, which were triangulated with the quantitative findings and provided explanatory context. The overall study design is shown in **Figure 1**.

**Figure 1:**
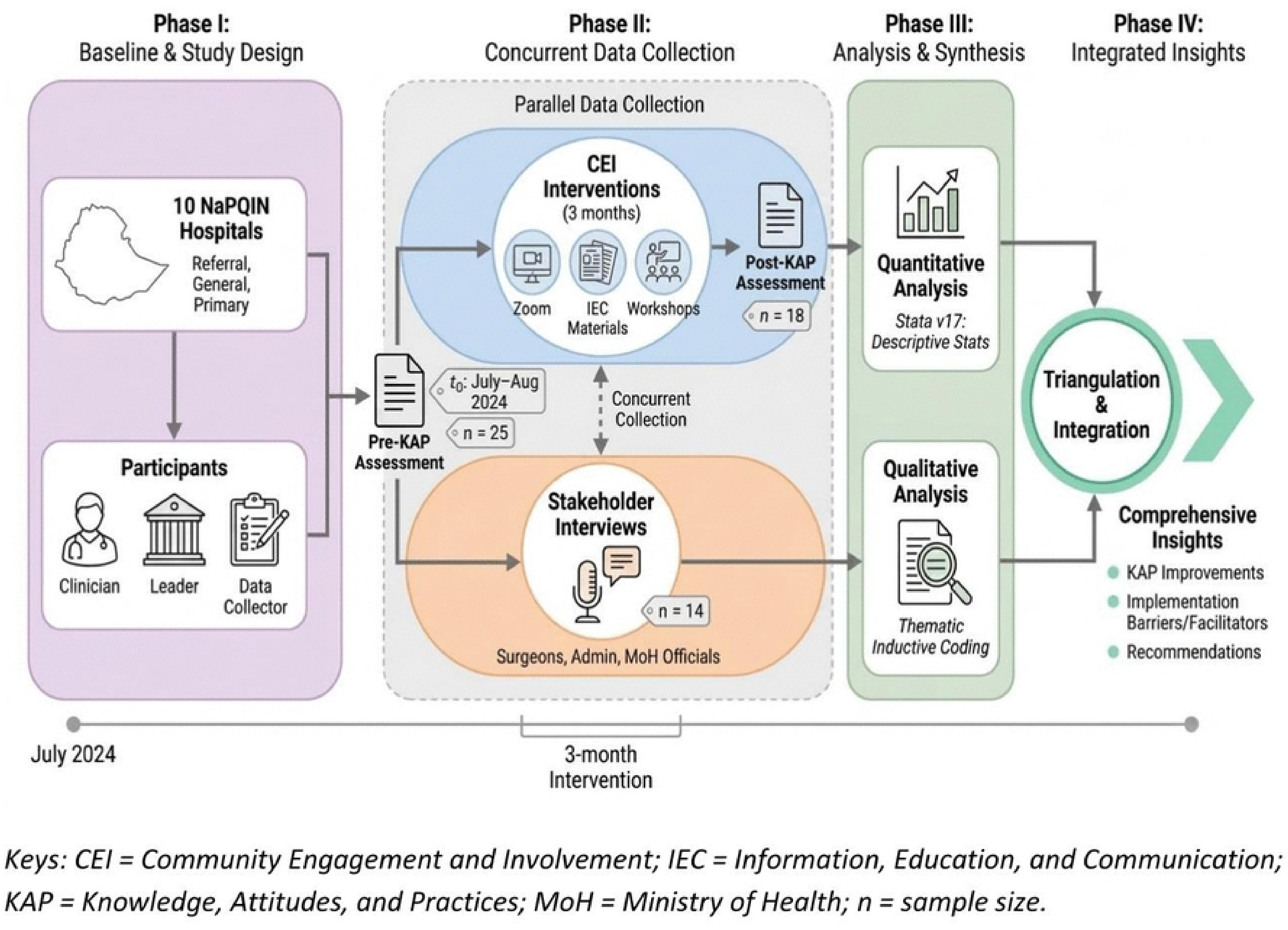
Overview of the convergent parallel mixed-methods study design. Keys: CEI= Community Engagement and Involvement; IEC= Information, Education, and Communication; KAP= Knowledge, Attitudes, and Practices; MoH= Ministry ofHealth; n= sample size.

#### Theme 1: Critical Importance and recognized value of perioperative data

All participants viewed perioperative data as essential to quality improvement, evidence-based practice, resource allocation, and policymaking. Surgeons emphasized personal clinical benefits, monitoring complication rates, and refining operative techniques, while administrators underscored its utility for theatre efficiency and local QI planning. Ministry of Health officials highlighted national KPI monitoring, including surgical volume and surgical site infection surveillance, as well as strategic planning and resource advocacy. One surgeon articulated the foundational rationale succinctly: “I need to know the real outcome… to decrease morbidity and mortality of all my patients, I need data.” Perioperative data were consistently framed as revealing the ‘real scenario’ of surgical activity and as enabling improvements in patient outcomes.

#### Theme 2: Barriers to high-quality data collection and registry implementation

Participants identified several interlocking barriers:

- Paper-based registers are the dominant documentation modality, associated with loss, illegibility, incompleteness, transcription errors, missing charts, and difficulty with retrospective capture after early patient discharge
- High clinical workload driving missed or hurried entries by surgeons, residents, and nurses, with documentation perceived as competing with direct patient care
- Knowledge and classification gaps, including misclassification of cases (e.g., elective versus emergency) and limited understanding of data importance at frontline clinical levels
- Resource constraints encompassing limited dedicated data staff, unreliable internet connectivity, slow electronic medical record rollout, inadequate training budgets, and regional security disruptions. Variable clinicians’ buy-in was also noted, with some viewing documentation as peripheral to their core clinical duties

#### Theme 3: Facilitators and strategies for successful implementation

Despite these barriers, participants identified several effective enablers:

- Regular mentorship and supervision, including twice-weekly data completeness checks with immediate corrective feedback, were credited with substantially improving data quality: ‘We check twice a week… Now there is a minimum problem’
- Stakeholder orientation and real-time demonstration of registry value, through dashboard outputs and QI feedback sessions, increased clinician ownership and compliance with documentation requirements
- Hybrid paper-digital systems: offline mobile data entry at the point of care reduced data loss and supported prospective rather than retrospective capture
- Structured processes including standardized source documents, cross-verification between operating theatre logs and registry entries, and designated data champions at each site
- Persistence and iterative adaptation, adding dedicated staff, adjusting rosters, and demonstrating tangible local uses of registry data, were critical to sustaining momentum through implementation challenges

#### Theme 4: Recommendations for other similar settings

Participants articulated three core transferable recommendations:

- Establish clear, standardized source documentation capturing all essential clinical indicators from the outset of implementation, rather than retrospectively introducing data standards
- Priorities repeated training, ongoing mentorship, and targeted awareness campaigns among frontline staff and institutional leaders to build technical skills and shift attitudes over time
- Secure early and sustained stakeholder buy-in across all levels, from operating theatre staff to Ministry of Health policymakers, and ensure adequate resourcing in personnel, technology, and budget from program inception

Additional guidance included: data entry during the operative period rather than retrospectively; integration with existing national Health Management Information System (HMIS) platforms; and emphasis on the local clinical and managerial value of data rather than framing data collection solely as a reporting obligation to external parties.

## Discussion

This mixed-methods evaluation suggests that structured community engagement and involvement supported the early implementation of NaPQIN by improving stakeholder knowledge and attitudes, with smaller gains in practice. At the same time, persistent workflow challenges and limited progress in benchmarking indicate that early implementation remained uneven. The study is distinctive in evaluating a structured CEI intervention within a national perioperative quality network in a low-resource setting, an area that has been little explored in the published literature.

These gains were most evident in knowledge. Knowledge improved across most assessed domains, particularly in understanding the primary purpose of surgical registries and recognizing what a perioperative registry is not. This pattern is important because these conceptual domains often undermine early implementation when frontline teams are uncertain about why data are collected and how to use them. Our findings are consistent with prior work showing that successful perioperative registries in low-resource settings depend on local stakeholder engagement, repeated orientation, and context-specific training (3,5,10). They also align with emerging implementation evidence from Ethiopia and Zambia showing that low-cost perioperative registries become more feasible and usable when iterative training and local ownership are built into design and rollout (4,5).

The qualitative findings reinforce this interpretation. Participants linked poor documentation, inconsistent data capture, and uncertainty about the registry’s value to a limited understanding of the registry’s purpose. That convergence between the quantitative and qualitative results suggests that the CEI program was responsive to frontline implementation needs rather than delivering generic awareness content. The few domains in which knowledge declined slightly, such as advocacy methods and broader program goals, suggest that refresher sessions may be needed to reinforce system-level concepts while implementation remains focused on immediate operational tasks (3,5,10).

The same pattern was seen in attitudes. Attitudes improved consistently across all measured domains, with the largest gains in overall satisfaction with NaPQIN, perceived contribution of CEI to sustainability, and the importance of community engagement for quality improvement. These patterns suggest that participants came to view CEI not as a parallel activity, but as part of how a registry becomes sustainable and useful. This interpretation is supported by broader CEI and patient-public engagement literature from sub-Saharan Africa, which argues that engagement is most effective when it moves beyond consultation toward meaningful participation, shared ownership, and integration into routine health-system processes (7,8,11,12).

However, this positive shift was not without tension. The reverse-scored item on workflow integration moved in an unfavorable direction, indicating that enthusiasm for the registry can coexist with continuing concern about documentation burden and workflow disruption. This is an important implementation message. In settings where staffing, time, and digital systems are limited, positive attitudes alone are unlikely to produce sustained behavioral change unless workflow barriers are addressed directly. The finding also echoes implementation reviews showing that training and stakeholder buy-in are necessary but insufficient unless they are accompanied by practical system support, iterative feedback, and enabling organizational conditions (13,14).

These constraints are also reflected in the practice findings. Practice-level change was more modest than the shifts in knowledge and attitudes, which is expected during early implementation. The largest gains were in clinicians’ use of the registry in daily work and in the formalization of CEI within quality improvement activities, suggesting that the intervention helped move the registry from a project concept toward routine practice. However, the smaller change in use of registry data for formal quality improvement projects and the decline in benchmarking use show that data collection matures before data use. This pattern is consistent with the perioperative implementation literature, in which successful scale-up often relies on training, stakeholder relationships, and evaluative feedback, but comparatively fewer programs report strong benchmarking or data-to-action mechanisms at scale (3,13,14).

The qualitative findings again help explain this gap. Participants described paper-based documentation, high workload, limited internet access, variable buy-in, and shortages of dedicated personnel as persistent constraints. They also highlighted the strategies that appeared to work: repeated mentorship, designated data champions, simple documentation tools, and opportunities to see how local data could inform action. Similar barriers and facilitators have been reported in perioperative registry implementation studies from Ethiopia and Zambia, where data completeness, capture, and usability improved only after local processes were refined and frontline teams were supported through iterative adaptation (4,5).

These findings also have implications for perioperative quality improvement in low- and middle-income countries. National registries are often discussed in technical terms, but this study shows that their sustainability also depends on social and organizational conditions: whether stakeholders understand the registry’s purpose, trust its value, receive feedback, and see a role for themselves in its success. In that sense, CEI should be considered implementation infrastructure rather than an optional add-on. For NaPQIN and similar networks, this means embedding repeated orientation, co-design with frontline users, supportive supervision, and patient or community perspectives into routine program delivery (7,8,11,12,14,15).

This study has several limitations. The pre- and post-intervention samples were small and not matched at the individual level, which limits causal inference and raises the possibility of selection or retention bias. Self-reported hospital membership of NaPQIN was low (40% before and 50% after the intervention) even though all respondents were sampled from NaPQIN hospitals. This may reflect limited awareness of formal membership among respondents who were not directly involved in registry activities, rather than non-membership. We did not verify membership status independently, and this item should be interpreted with caution. The absence of a comparison group means that changes cannot be attributed solely to the CEI program, as concurrent NaPQIN activities or broader system developments may also have contributed. The qualitative sample was intentionally diverse but relatively small, and the findings may not transfer fully to settings without an existing multi-hospital network or similar leadership support. Finally, follow-up was limited to the immediate post-intervention period, so the durability of the observed gains remains uncertain. Future studies should use larger samples, longer follow-up, and comparative designs to assess whether early gains in knowledge and attitudes translate into sustained improvements in registry use and patient-centered quality improvement (3,13,14).

## Conclusions

This study suggests that a structured community engagement and involvement program improved stakeholders’ knowledge of perioperative registry functions and strengthened positive attitudes toward NaPQIN across measured domains, with emerging gains in practice. The convergence of quantitative and qualitative findings suggests that CEI is not merely a supportive add-on but a core mechanism for strengthening knowledge, attitudes, and practice within a national perioperative quality network. In similar low- and middle-income country settings, CEI should be embedded as a recurring, adequately resourced program component. Future work should strengthen benchmarking capacity, co-design registry workflows with frontline staff, improve digital infrastructure, and evaluate long-term sustainability with larger sample sizes, matched comparison sites, and longitudinal follow-up.

## Data Availability

Availability of data and materials All relevant data are within the manuscript and its Supporting Information file (S1 File), which contains the de-identified aggregate survey data underlying Tables 1 to 4. The interview transcripts cannot be shared because they contain potentially identifying information, and public deposition was not covered by the ethical approval (Debre Berhan University Asrat Woldeyes Health Science Campus IRB, Protocol No. IRB-095). The interview findings are presented as themes and quotations in the Results.

## Supporting information

S1 File. De-identified aggregate survey data (counts and percentages for each item, pre- and post-intervention).

### Funding

This research was funded by the NIHR (NIHR133850) using UK international development funding from the UK Government to support global health research. The views expressed in this publication are those of the authors and not necessarily those of the NIHR or the UK government.

### Authors’ contributions

All authors contributed to the conception, design, implementation, data collection, analysis, and manuscript preparation. All authors reviewed and approved the final manuscript.

### Availability of data and materials

All relevant data are within the manuscript and its Supporting Information file (S1 File), which contains the de-identified aggregate survey data underlying Tables 1 to 4. The interview transcripts cannot be shared because they contain potentially identifying information, and public deposition was not covered by the ethical approval (Debre Berhan University Asrat Woldeyes Health Science Campus IRB, Protocol No. IRB-095). The interview findings are presented as themes and quotations in the Results.

